# Large-scale genome-wide analyses of proteomic data identifies that sex hormones affect plasma glycodelin levels

**DOI:** 10.64898/2026.03.06.26347586

**Authors:** Sophia McDowell, Robin N. Beaumont, Harry Green, Rebecca Kingdom, Marina Vabistsevits, Julia K. Prague, Anna Murray, Jessica Tyrrell, Katherine S. Ruth

**Author notes:** Correspondence: Katherine S. Ruth.

## Abstract

**Study question:** How is glycodelin, a glycoprotein secreted by reproductive tissues, causally related to reproductive diseases and traits?

**Summary answer:** We present evidence for a causal role of sex hormones in determining glycodelin levels, but limited evidence that glycodelin subsequently causally impacts reproductive traits.

**What is known already:** Glycodelin is expressed in female and male reproductive tissues and has four glycoforms (-A, -C, -F and -S), with the glycosylation pattern determining its function. Differences in the levels of glycodelin are associated with reproductive traits, including fertility, endometriosis, preeclampsia, and female-specific malignancies.

**Study design, size, duration:** We used cross-sectional data from the UK Biobank to investigate relationships between glycodelin and reproductive-related traits in men and women by performing genome-wide association studies (GWAS) and Mendelian randomization (MR) analyses.

**Participants/materials, setting, methods:** We included individuals of European genetic ancestry aged 40–69 in 2006–2010, with genetic data in the UK Biobank v3 release. We performed GWAS of glycodelin levels in 46,468 people, stratified by sex (21,368 men and 25,100 women) and menopause status (6,409 pre- and 18,691 post-menopausal women). We tested bidirectional casual associations between glycodelin levels and 19 reproductive-related traits using one- and two-sample MR analyses.

**Main results and the role of chance:** Nine genetic signals reached genome-wide significance (P<5×10^-8^) across the glycodelin phenotypes. A known genetic signal (rs9409964) near the *PAEP* gene, which encodes glycodelin, was most strongly associated (*P*<3×10^-80^ across all phenotypes), and had heterogeneous effects (effect (SD) per A allele of 1.31 in men vs 0.60 in women, and 0.4 in pre- vs 0.9 in post-menopausal women). Higher serum concentrations of bioavailable testosterone raised glycodelin in men (effect = 0.14 SD, IVW *P*=4.1×10^-13^), while effects in women depended on menopause status (pre-menopausal effect = -0.16 SD, IVW *P*=3.6×10^-3^; post-menopausal effect = 0.10 SD, IVW *P*=5.9×10^-4^). There was no strong evidence that differences in glycodelin levels were caused by, or were the cause of, other reproductive-related traits.

**Limitations, reasons for caution:** Proteomic measurements of glycodelin did not differentiate between glycoforms and were derived from blood and might not reflect levels in reproductive tissues. The sample size for the pre-menopausal GWAS was modest, reducing our power to detect relationships with reproductive conditions. Genetic instruments are assumed to be proxies for average lifelong exposure, which does not reflect variation in hormones and biomarkers over lifetime.

**Wider implications of the findings:** We suggest that reported associations of glycodelin with reproductive conditions are likely to result from the effects of sex hormones rather than being directly causal. These findings may help reconcile previously conflicting associations between glycodelin and reproductive traits.

## Introduction

Glycodelin, or progesterone-associated endometrial protein, plays an important role in fertilisation and pregnancy, and has been implicated in various reproductive conditions (Seppälä *et al*., 2002; Lapid and Sharon, 2006; Uchida *et al*., 2013). Glycodelin is expressed in secretory and decidualised endometrium, bone marrow, ovaries, fallopian tubes and seminal vesicles and has four glycoforms (-A, -C, -F and -S), with the glycosylation pattern determining its function (Seppälä *et al*., 2002; Lapid and Sharon, 2006; Uchida *et al*., 2013). In men, glycodelin-S is produced by the seminal vesicles and binds to sperm to maintain an inactive state. In the female reproductive system, glycodelin levels are regulated by progesterone and are involved in modulating the fertilisation of the oocyte by the sperm. Glycodelin-A is produced by the secretory endometrium and has roles in preventing fertilisation and supporting maternal immunosuppression during implantation. Glycodelin-F is found in granulosa cells and inhibits sperm activation. Finally, glycodelin-C is produced by the cumulus cells surrounding the oocyte and stimulates sperm binding (Lapid and Sharon, 2006; Chiu *et al*., 2007; Uchida *et al*., 2013; Yadav *et al*., 2023).

Differences in glycodelin levels are associated with reproductive traits, suggesting that glycodelin could be used as a biomarker for endometrial receptivity, endometriosis and reproductive cancers (Suzuki *et al*., 2000; Lapid and Sharon, 2006; Seppälä *et al*., 2009; Tsviliana *et al*., 2010; Scholz *et al*., 2012; Kocbek *et al*., 2013; Wang *et al*., 2013; Cui *et al*., 2017; Pant *et al*., 2023; Sun *et al*., 2023). Lowered serum glycodelin levels are associated with infertility or recurrent miscarriage (Suzuki *et al*., 2000), and raised serum levels with the presence and severity of preeclampsia (Dundar *et al*., 2018). In women with endometriosis, glycodelin levels are elevated in serum and/or peritoneal fluid (Drosdzol-Cop and Skrzypulec-Plinta, 2012; Kocbek *et al*., 2013; Wang *et al*., 2013; Mosbah *et al*., 2016) and glycodelin expression is downregulated in secretory-phase endometrium (Burney *et al*., 2007). However, such studies are unable to confirm whether glycodelin causes, is caused by, or is merely correlated with reproductive traits.

The recent availability of large-scale proteomics assays in population cohorts with genomic data provides an opportunity to explore aetiological relationships of glycodelin with health and disease. A recent proteome-wide study in 54,219 people in UK Biobank identified a decline in plasma levels of glycodelin at menopause in women, while in men glycodelin levels increased steadily with age (Supplementary Figure 1) (Sun *et al*., 2023). Sun et al (2023) identified two genetic variants associated with glycodelin levels, with further analyses identifying a gene-by-environment interaction between body weight and glycodelin levels that differed in pre- and post-menopausal women (Hillary *et al*., 2024). Such genetic variants can be used as instruments in Mendelian randomization (MR) analysis to test associations between genetically-determined epidemiological exposures and outcomes (Davey Smith and Ebrahim, 2003). Since genotype should not be affected by risk factors over life, MR methods are robust to confounding and reverse causality and provide evidence of causal relationships. Therefore, MR can be used to investigate the role of glycodelin in reproductive health, and the results of these previous genetic studies suggest that genetic instruments for glycodelin might be specific to sex and menopause status.

Using proteomic data from UK Biobank, we conducted stratified genome-wide analyses of glycodelin levels in up to 46,468 people. Using results from our genetic association analyses, we tested the causality and direction of relationships between glycodelin and 19 reproductive-related conditions by performing MR analyses. We identified little evidence of relationships between glycodelin levels and reproductive health implicated by observational studies, but infer likely causal associations with sex hormone levels, which differ by sex and menopause status.

## Methods

### Phenotype definitions in UK Biobank

The UK Biobank is a population study with 500,000 participants aged between 40-69 years from across the UK (Allen *et al*., 2024). The study was approved by the North West Multi-centre Research Ethics Committee as a research tissue bank. All participants provided written informed consent, and all study procedures were performed in accordance with the World Medical Association Declaration of Helsinki ethical principles for medical research.

Phenotypes for our GWASs were glycodelin levels generated by Olink as part of the UK Biobank Pharma Proteomics Project. Proteomic profiling of 2,923 proteins in 54,219 UK Biobank participants was conducted on blood plasma samples at baseline using antibody-based Olink proximity extension assays. After quality control procedures, protein levels were normalized before release. Further details of the assays, data processing and quality control are discussed elsewhere (Sun *et al*., 2023).

Menopause status at baseline visit was derived from self-reported questionnaire data based on variables relating to age at menopause, hysterectomy, oophorectomy and hormone replacement therapy use (Ruth *et al*., 2016). We included women who had gone through surgical and natural menopause in the post-menopausal phenotype.

Sex hormone binding globulin (SHBG), total testosterone and oestradiol levels were measured in serum at baseline, as part of a panel of 34 biomarkers. We also calculated bioavailable testosterone, as detailed in a prior publication (Ruth *et al*., 2020).

### Genome-wide association analyses

We performed genome-wide association study (GWAS) analyses of glycodelin levels in 46,468 people in UK Biobank, further stratified by sex (21,368 men and 25,100 women) and menopause status (6,409 pre-menopausal and 18,691 post-menopausal women), resulting in five GWAS in total. We included individuals identified as of European genetic ancestry who had matching genetically-inferred and self-reported sex (Tyrrell *et al*., 2019). We used genetic data from the ‘V3’ release of UK Biobank, which contains the full set of Haplotype Reference Consortium and 1000 Genomes imputed variants. Details of central genotyping, quality control and imputation are described in full elsewhere (Bycroft *et al*., 2018). Association testing was performed using REGENIE (v3.3) (Mbatchou *et al*., 2021) and was based on an additive model with adjustment for age, genotyping chip/batch, recruitment centre, 40 ancestral principal components, sex (sex combined analysis only) and type of menopause (female analysis only). Glycodelin levels were transformed by inverse rank normalization prior to analysis. For quality control purposes, we restricted genetic variants in our analyses to have both a minor allele frequency (MAF) > 0.1% and minor allele count > 50, and imputation quality > 0.4. We considered genome-wide significant signals to be genetic variants with *P*<5×10^-8^ that were more than 500kb from the next such variant. Signals across the phenotypes that were within 500kb of each other and in strong linkage disequilibrium (LD) (r^2^ > 0.8) were considered to be in the same locus and where there were multiple signals at a locus, we considered the variant from the ‘All’ phenotype to be the ‘lead’ variant for that locus.

To estimate the magnitude of effects of the genetic signals in the untransformed normalized protein expression (NPX) values, we multiplied effect estimates from our GWAS by the standard deviation of the untransformed phenotype (Supplementary Table 1). Due to a low minor allele count, variant rs184353251 was not analysed in the GWAS of pre-menopausal glycodelin levels. To estimate an effect size for rs184353251 in pre-menopausal women, we extracted the genotype from the bulk imputed files using the R package ‘ukbrapR’ (https://lcpilling.github.io/ukbrapR) and performed linear regression of inverse rank normalized glycodelin in 5,395 unrelated women (Tyrrell *et al*., 2019) adjusted for the same covariates as in the GWAS. Differences in effect sizes were compared between men and women, and pre- and post-menopausal women using a two-sample z-test of differences in means.

Lambda-genomic control was calculated for each phenotype to assess population stratification. Manhattan, quantile−quantile and locus plots were generated for each GWAS in R (v4.4.0) using the package ‘topr’ (v 2.0.2) (Juliusdottir, 2023). We annotated the GWAS signals with nearby genes using PLINK (v1.9) (Purcell *et al*., 2007) and with genomic consequence using Ensembl Variant Effect Predictor (McLaren *et al*., 2016). FUMA was also used for its SNP2GENE module to identify eQTLs in reproductive tissues (uterus, testis, ovary, vagina and prostate) from the Genotype-Tissue Expression (GTEx) database (v8) and GWAS catalogue associations for lead variants and proxies with r2>0.8 (Watanabe *et al*., 2017). LD information was for European ancestry based on TOPMed whole genome sequencing data and was obtained using the TopLD tool (Huang *et al*., 2022).

### Identification of genetic instruments for MR

We identified genetic instruments to use as genetically-determined exposures in our MR analyses. MR assumes that: (i) the instrument must be associated with the exposure; (ii) the instrument must be independent of confounders between the exposure and outcome; (iii) there must be no independent pathway between the instrument and the outcome other than via the exposure. All genetic signals included in our instruments were robustly associated in GWAS at *P*<5×10^-8^, satisfying the first assumption.

For the exposure of glycodelin levels, we used a genetic instrument based on a single variant in *cis* with the *PAEP* gene that we identified from our GWAS (rs9409964). Variants in *cis* with a gene are likely to regulate protein levels directly, making them less susceptible to violating the MR assumptions.

We identified genetic instruments for 16 exposure traits with relevance to reproductive health (‘reproductive-related traits’). These were traits with large GWAS available that have been previously linked to glycodelin in observational studies or that were related to reproductive lifespan, reproductive conditions, hormone sensitive cancers, sex hormones, fertility/pregnancy, or adiposity (Supplementary Table 2).

They included age at menarche and age at natural menopause (ANM); endometriosis, polycystic ovary syndrome (PCOS) and uterine leiomyomata (fibroids); female breast cancer, endometrial cancer and ovarian cancer; bioavailable testosterone level, male oestradiol level, total testosterone level and SHBG level; pre-eclampsia and reproductive success; and BMI and waist:hip ratio (WHR). Such genetic instruments included 10 or more genetic signals to enable robust MR estimates to be generated. All genetic instruments had an F-statistic>10 (Supplementary Table 3) indicating sufficient power in the MR analyses (Sanderson *et al*., 2021). Where possible, we used sex-specific effect estimates. Since considerable overlap exists in the underlying genetics of sex hormone regulation, we used genetic instruments largely specific to individual hormones in each sex to separate their individual contributions. These were previously identified by clustering methods and as such, we refer to these as ‘clustered’ instruments (Ruth *et al*., 2020). Female oestradiol level was not included due to a lack of a robust genetic instrument (Ruth *et al*., 2020).

### MR analyses

We used the TwoSampleMR R package (v0.6.15) (Hemani *et al*., 2018) to perform two-sample MR, which uses summary-level GWAS data to estimate the effects of an exposure on an outcome (Davies *et al*., 2018). We tested causal associations between the exposure of glycodelin levels with 18 reproductive-related traits as the outcome by calculating the Wald ratio. As outcomes, we included reproductive-related traits (as described previously) with publicly-available GWAS summary statistics and, where possible, sex-specific effect estimates (Supplementary Table 2). The *cis* variant was not present in the BMI outcome GWAS; therefore, a proxy SNP (rs697449, r^2^ = 0.998) was used as a genetic instrument. To identify significant associations, we used a Bonferroni corrected p-value threshold of *P*<1.4×10^-3^ (0.05/35 tests carried out in the ‘all’, ‘male’ and ‘female’ phenotypes).

For the analyses of reproductive-related traits as exposure on glycodelin levels as outcome, we performed inverse-variance weighted (IVW) MR. As sensitivity analyses, we performed simple mode MR, weighted mode MR, weighted median MR and MR Egger, which are more robust to pleiotropy (Bowden *et al*., 2016b, 2016a). For binary exposures, we estimated the effect on glycodelin levels per doubling of the probability of the exposure (Burgess and Labrecque, 2018). Associations with IVW MR p-values below a Bonferroni-corrected significance threshold of *P*<1.8×10^-3^ were considered significant (0.05/28 tests in the ‘all’, ‘male’ and ‘female’ groups), provided that results of the sensitivity analyses were directionally consistent.

Additionally, as a sensitivity analysis, we carried out one-sample MR in UK Biobank for reproductive-related exposure traits where the published GWAS were in UK Biobank only (sex hormones) or had a large proportion of UK Biobank participants (age at natural menopause). A strength of one-sample MR is that the exposure and outcome groups are from the same underlying population, an assumption of MR (Burgess *et al*., 2023). One-sample MR was performed using the two-stage least-squares regression estimator method. We predicted individuals’ sex hormone level or age at natural menopause based on their genotype by calculating a genetic score, and then regressed the outcome of glycodelin levels against these predicted values. Genetic scores were generated using a function detailed in the GitHub repository GRS-Nexus (https://github.com/hdg204/GRS-Nexus). Results from the two-sample IVW were taken as our primary analyses since only a subset of the UK Biobank had proteomic measures available, restricting the sample size for one-sample MR.

This study is reported as per the guidelines for strengthening the reporting of Mendelian randomization studies (STROBE-MR) (Skrivankova *et al*., 2021).

## Results

### Nine genetic loci associated with glycodelin levels

Our GWAS identified nine independent genetic loci across all phenotypes that reached genome-wide significance (*P*<5×10^-8^) (Table 1, Supplementary Figure 3, Supplementary Table 4) of which seven were associated with glycodelin levels in the sex-combined (‘All’) analysis. We replicated two signals (near the genes *PAEP* and *ZNF565*) previously reported for glycodelin in UK Biobank, and identified six signals that passed genome-wide significance in our study that did not pass *P*<5×10^-8^ in a previous study (Sun *et al*., 2023). Consistent with the previous UK Biobank analysis, the genetic variant rs9409964, which is 13kb away from the *PAEP* gene that encodes the glycodelin protein, was most strongly associated across all phenotypes (*P*<3×10^-80^ for all phenotypes) (Supplementary Table 4).

**Table 1:** Independent genetic signals identified from GWAS of glycodelin levels. Results are from five GWAS of glycodelin levels in males and females combined (‘All’), males and females separately, and in pre- and post-menopausal females. Effect shown is for the glycodelin raising allele.

| Locus | Lead variant | Chr_Pos_EA_OA | Phenotype | EAF | Effect | SE | P | Nearest protein coding gene (distance) |
| --- | --- | --- | --- | --- | --- | --- | --- | --- |
| 1 | **rs1130866** | 2:85893741_G_A | **All** | **0.484** | **0.038** | **0.005** | **2.24E-17** | *SFTPB* (0kb) |
|  |  |  | Females | 0.481 | 0.045 | 0.006 | 2.00E-13 |  |
|  |  |  | Post-menopause | 0.481 | 0.065 | 0.009 | 6.03E-14 |  |
| 2 | **rs62449498** | 7:22778528_C_A | **All** | **0.491** | **0.026** | **0.005** | **1.62E-08** | *IL6* (+6.91kb) |
| 3 | **rs9409964** | 9:138471928_A_G | **All** | 0.188 | **0.869** | **0.007** | P<1E-50 | *PAEP* (+13.31kb) |
|  |  |  | Females | 0.188 | 0.604 | 0.009 | P<1E-50 |  |
|  |  |  | Males | 0.187 | 1.309 | 0.012 | P<1E-50 |  |
|  |  |  | Pre-menopause | 0.190 | 0.391 | 0.021 | P<1E-50 |  |
|  |  |  | Post-menopause | 0.188 | 0.905 | 0.013 | P<1E-50 |  |
| 4 | **rs1265847** | 10:28929686_A_G | **All** | **0.263** | **0.032** | **0.005** | **4.27E-10** | *WAC* (+17.64kb) |
|  | rs2532741 | 10:28863808_G_C | Males | 0.251 | 0.057 | 0.007 | 1.95E-14 |  |
| 5 | **rs58118673** | 10:49787836_CT_C | **Pre-menopause** | **0.955** | **0.216** | **0.039** | **2.40E-08** | *ARHGAP22* (0kb) |
| 6 | **rs78435901** | 16:79218585_C_G | **All** | 0.948 | **0.058** | **0.010** | 1.05E-08 | *WWOX* (0kb) |
| 7 | **rs184353251** | 17:65641428_A_G | **Post-menopause** | **0.002** | **0.526** | **0.092** | **1.12E-08** | *PITPNC1* (0kb) |
| 8 | **rs7249719** | 19:36681413_A_G | **All** | **0.478** | **0.045** | **0.005** | **1.10E-23** | ZNF565 (0kb) |
|  | rs10420462 | 19:36666803_C_T | Females | 0.470 | 0.043 | 0.006 | 4.90E-12 |  |
|  | rs1974843 | 19:36680555_T_A | Males | 0.482 | 0.054 | 0.006 | 4.07E-17 |  |
|  | rs10401633 | 19:36659354_C_T | Post-menopause | 0.483 | 0.056 | 0.009 | 5.89E-11 |  |
| 9 | **rs35106244** | 19:49203829_T_C | **All** | **0.464** | **0.029** | **0.005** | **5.62E-10** | FUT2 (0kb) |

*Positions are in b37. Effect sizes are in SD of the inverse normalised phenotype. Where there were multiple signals at a locus, we considered the variant* *from the 'All' phenotype the 'lead' variant for that locus (highlighted in bold). EAF: effect allele frequency; CI: confidence interval; Chr\_Pos\_EA\_OA:* *chromosome, position, effect allele and other (non-effect) allele; SE: standard error.*

### Genetic basis differs by sex and menopause status

The effects of rs9409964 (near *PAEP*) differed by sex and menopause status, though the A allele was associated with raised glycodelin levels across all five phenotypes (Table 1 and Figure 1). Signal rs9409964 had a greater effect on glycodelin levels in men (effect = 1.31 SD (95% CI 1.28,1.33 SD) per A allele (AF=0.188), P<1x10^-50^) than women (effect = 0.60 SD (95% CI 0.59, 0.62 SD), P<1x10^-50^; *P*_difference_<1×10^-50^) (Supplementary Table 5). In women, the effect of rs9409964 was approximately twice as large in post-menopausal compared with pre-menopausal individuals (0.91 vs 0.40 SD per A allele respectively; *P_difference_*<1×10^-50^). However, when converted to raw units of normalized protein expression (NPX), the effect of rs9409964 was similar in both groups (0.8 NPX per A allele for both; *P_difference_*=0.88), reflecting greater variation in glycodelin levels in pre-versus post-menopausal women (SD=2.04 vs 0.89 respectively) (Supplementary Figure 2 and Supplementary Table 1). Although rs9409964 was not an eQTL in reproductive tissues (Supplementary Table 4), the variant was an eQTL for *PAEP* in other tissues (P=3.4×10^-33^ for minor salivary gland), as well as *OBP2A*, *LINC01502* and *MRPS2* (P<5×10^-8^ for all), with the glycodelin raising allele associated with raised expression.

**Figure 1.**
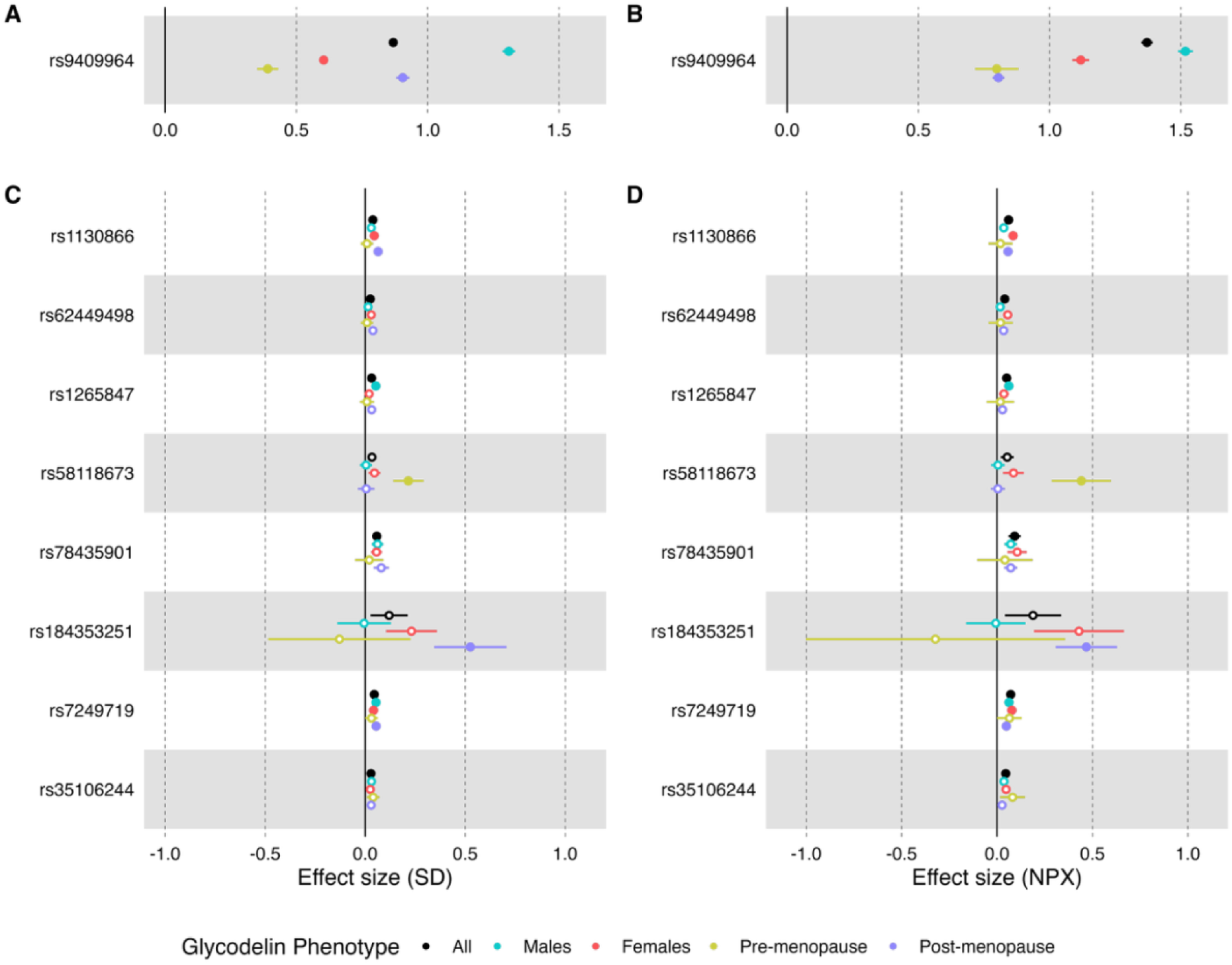
Effect size of genetic associations with glycodelin levels by sex and menopause status. Effect size of rs9409964 (nearest signal to PAEP) in (A) SD and (B) scaled to NPX. Effect sizes of additional variants with sex- or menopause-specific effects in (C) SD and (D) scaled to NPX. *Solid points indicate genome-wide significant associations (P < 5 x 10^-8^). Error bars show 95% confidence intervals*.

Two signals only reached genome-wide significance when analyses were stratified by menopause status (Figure 1 and Supplementary Table 4). Signal rs58118673 (intronic in *ARHGAP22*) had a stronger effect in pre-menopausal women (effect=0.22 SD (95% CI 0.14,0.29) per CT allele (AF=0.955), *P*=2.4×10^-8^), than post-menopausal women (effect=-0.01 SD (95% CI -0.04,0.05), *P*=0.81; *P_differenc_*_e_<1×10^-50^). In contrast, rs184353251 (intronic in *PITPNC1*), reached genome-wide significance in post-menopausal women (effect=0.53 SD (95% CI 0.35,0.71) per A allele (AF=0.002), *P*=1.1×10^-8^), but not pre-menopausal women (effect=-0.13 SD (95% CI -0.49,0.23), *P*=0.34; *P_difference_*=0.05).

Two further signals, rs1265847 and rs1130866, only reached significance in one sex though directions of effects were consistent. Signal rs1265847 (near *WAC*), had a stronger effect in men (effect=0.05 SD (95% CI 0.04,0.7) per A allele (AF=0.260), *P*=3.9×10^-13^) than women (effect=0.02 SD (95% CI 0.01,0.3), *P*=5.6×10^-3^; *P_difference_*=7.5×10^-4^). On chromosome 2, a single variant (rs1130866, a missense variant in *SFTPB*) passed genome-wide significance in women (effect=0.05 SD (95% CI 0.03,0.06) per G allele (AF=0.481), *P*=2.0×10^-13^), but was only nominally significant in men though effect estimates were similar (effect=0.03 SD (95% CI 0.02,0.04), *P*=2.8×10^-6^; *P_difference_*=0.10) (Figure 1, Supplementary Figure 3).

### Higher glycodelin does not change the risk of reproductive conditions

There was little evidence of causal associations between raised glycodelin, and the reproductive-related outcomes tested in the two-sample *cis*-MR analyses. There was some evidence for an association between glycodelin and three traits at *P*<0.05, though none passed a Bonferroni-corrected significance level (*P*=1.4×10^-3^) (Supplementary Table 6). Higher glycodelin levels lowered male BMI (effect=-0.01 SD per one-SD increase (95% CI -0.01,0.00), *P*=0.013) and raised female WHR (for WHR adjusted for BMI, effect=0.01 SD per one-SD increase (95% CI 0.00,0.02), *P*=0.039), with some suggestion of opposite effects in men and women. Elevated glycodelin levels increased the risk of ovarian cancer (OR=1.06 per one-SD increase (95% CI 1.00,1.12), *P*=0.045), with consistent effects before and after menopause (Figure 2), though glycodelin was not associated more generally with female hormone-related cancers.

**Figure 2.**
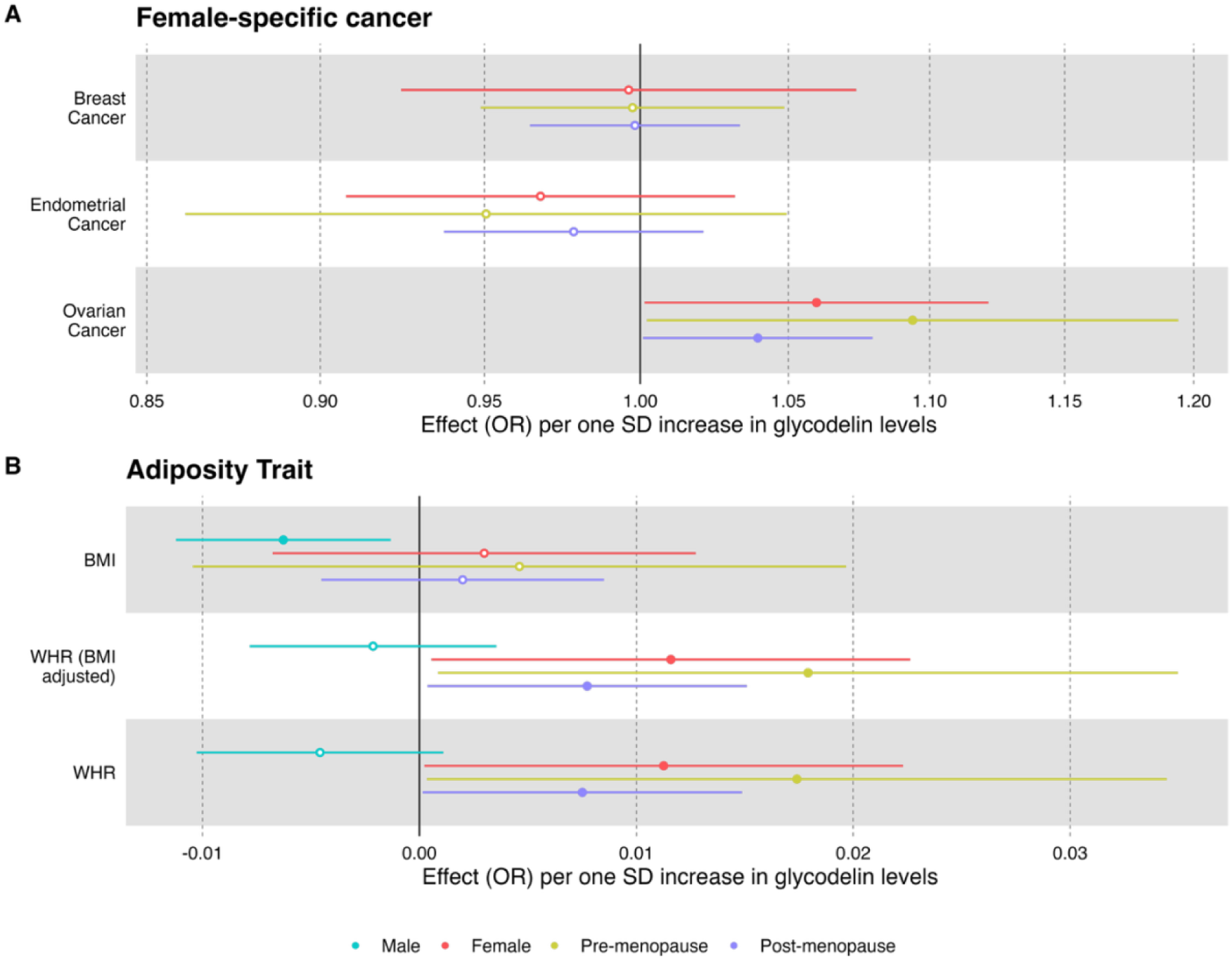
Results of Mendelian randomization analyses using genetic instruments showing effects of higher glycodelin by sex and menopause status on (A) female-specific cancer risk (B) adiposity-related measures. *Error bars show 95% confidence intervals around the point estimate from IVW analyses*.

### Glycodelin levels are affected by reproductive phenotypes

We identified robust causal associations between sex hormones and glycodelin (IVW *P*<1.8×10^-3^) with similar directions of effect in men and post-menopausal women, and opposite effects in pre-menopausal women (Supplementary Table 7, Supplementary Table 8, Figure 3). Higher bioavailable testosterone raised glycodelin in men (IVW effect=0.14 SD per one-SD increase (95% CI 0.11,0.18), *P*=4.1×10^-13^) and post-menopausal women (IVW effect=0.10 SD per one-ln(nmol/l) increase (95% CI 0.04,0.16), *P*=5.9×10^-4^), but had the opposite effect in pre-menopausal women (IVW effect=-0.16 SD per one-ln(nmol/l) increase (95% CI -0.27,-0.05), *P*=3.6×10^-3^). Higher testosterone raised glycodelin in men (IVW effect=0.18 SD per one-SD increase (95% CI 0.13,0.24), *P*=3.6×10^-11^) and post-menopausal women (IVW effect=0.065 SD per one-SD increase (95% CI 0.02, 0.11), *P*=5.1×10^-3^) but not pre-menopausal women (IVW effect=-0.07 SD per one-SD increase (95% CI -0.16,0.01), *P*=0.10). In contrast, higher SHBG raised glycodelin levels in pre-menopausal women (IVW effect=0.18 SD per one-SD increase (95% CI 0.05,0.30), *P*=6.5×10^-3^), but lowered glycodelin in post-menopausal women (IVW effect=-0.09 SD per one-SD increase (95% CI -0.15,-0.02), *P*=0.01).

**Figure 3.**
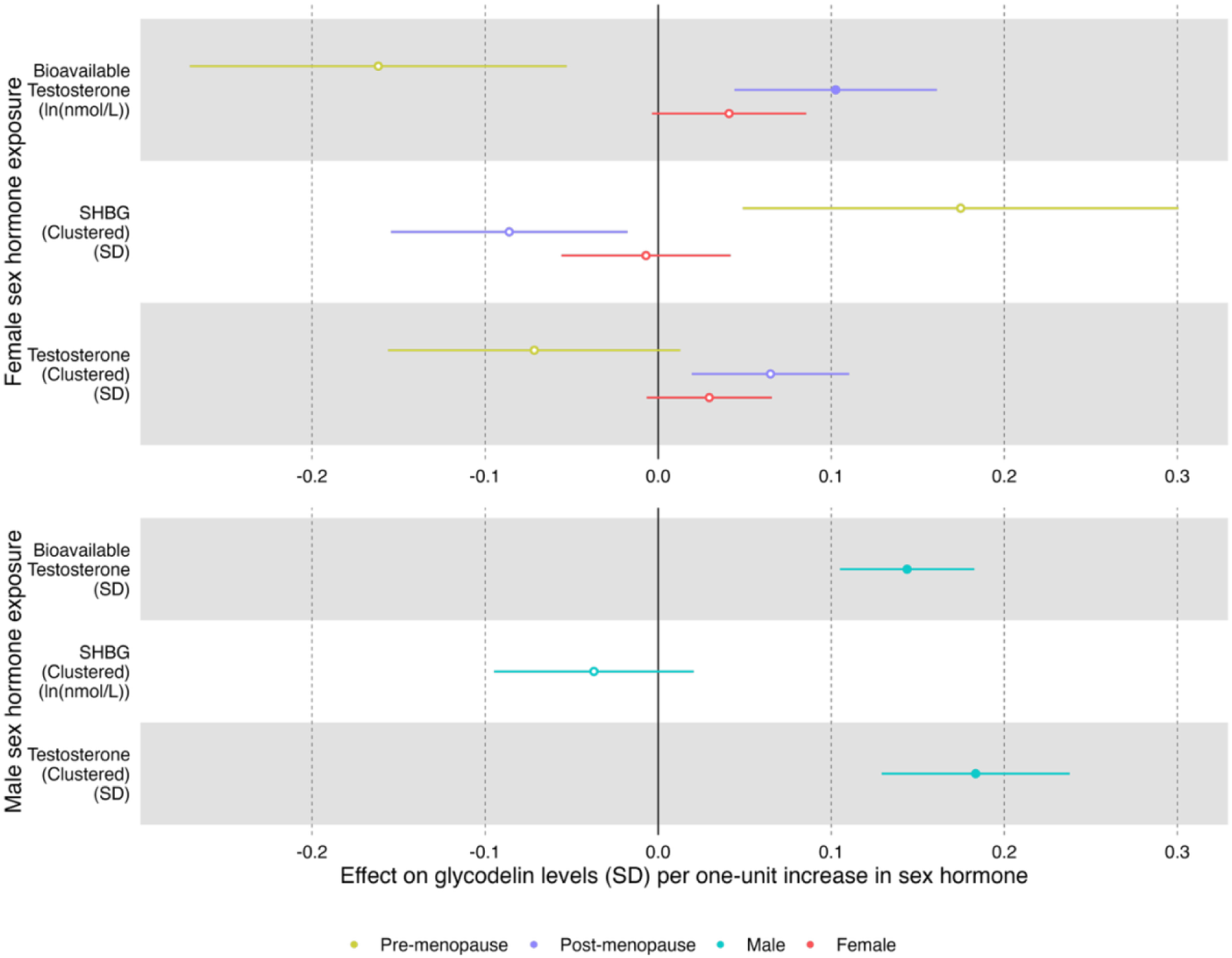
Results of Mendelian randomization analyses using genetic instruments showing effects of higher sex hormone concentrations on glycodelin levels by sex and menopause status. *Solid points indicate associations at P<1.8×10^-3^. Error bars show 95% confidence intervals around the point estimate from IVW analyses*.

Glycodelin levels were causally associated with additional reproductive-related traits. Consistent with the observational relationship between menopause and glycodelin, a later age of natural menopause raised glycodelin levels, with stronger effects in pre-menopausal women (0.06 SD per year (95% CI 0.04,0.08), *P*= 2.7×10^-11^) (Supplementary Table 7) than post-menopausal women (0.02 SD per year (95% CI 0.01,0.03), *P*= 3.2×10^-4^). There was a nominally significant relationship between later age at menarche and lower post-(but not pre-) menopausal glycodelin levels (-0.03 SD per year (95% CI: -0.05,-0.01), *P*=0.02). We further found that higher BMI lowers glycodelin levels in pre-menopausal women (-0.10 SD per one-SD increase (95% CI -0.18,-0.01), *P*=0.02), and though the effect was opposite in post-menopausal women, this was not supported by sensitivity analyses. Associations between higher odds of breast cancer and PCOS were not robust to sensitivity analyses, and other associations with reproductive cancers and other reproductive conditions did not reach nominal significance.

## Discussion

By performing genome-wide analyses stratified by sex and menopause status, we found that raised sex hormones have opposite effects on glycodelin levels before and after menopause. Having higher bioavailable testosterone lowered levels of glycodelin in pre-menopausal women but raised levels in post-menopausal women and in men. In women, glycodelin is produced by the endometrial epithelia, and the granulosa and cumulus cells of the ovary (which are not present after menopause), and seminal vesicles in men. Glycodelin is used experimentally to indicate the presence of secretory epithelial cells in the endometrium (Garcia-Alonso *et al*., 2021) and has been suggested as a biomarker for ovarian cancer (Cui *et al*., 2017) and endometriosis (Mosbah *et al*., 2016; Pant *et al*., 2023). In a previous MR study, higher bioavailable testosterone lowered the risk of endometriosis (McGrath *et al*., 2023), which when taken with our results, could explain observed relationships between elevated glycodelin and endometriosis (Kocbek *et al*., 2013; Wang *et al*., 2013; Pant *et al*., 2023). Other than sex hormone traits, we did not find any strong evidence to support a direct causal effect of reproductive conditions on glycodelin levels, or of differences in glycodelin levels being a risk factor for reproductive traits. However, the number of pre-menopausal women in our analyses was modest (n=6,409) and they were aged over 40 years which may have reduced our power to detect relationships with reproductive conditions. However, we did identify causal associations of raised testosterone, bioavailable testosterone and SHBG with glycodelin. Our results suggest that the suggested relevance of glycodelin as a potential biomarker for reproductive conditions is likely to be through its relationship with testosterone and SHBG and this may explain previous conflicting results.

Our study suggests that the effect of bioavailable testosterone on glycodelin is likely to be mediated through different biological pathways with different relative effects on testosterone and SHBG production depending on sex and menopause status. The amount of bioavailable testosterone (free testosterone plus that bound to albumin which is effectively available to cells and tissues) is reduced by higher levels of SHBG, and so to separate out these effects we used published genetic instruments that are more specific to these hormones (Ruth *et al*., 2020). The testosterone genetic instrument is composed of genetic variants with effects specific to testosterone. However, the SHBG genetic instrument has primary effects on SHBG but also secondary effects on bioavailable testosterone and testosterone, reflecting downstream effects of SHBG on these hormones. Our results suggest that effects of bioavailable testosterone on glycodelin are likely to be mediated through total testosterone in men, total testosterone and SHBG in post-menopausal women, and SHBG in pre-menopausal women and suggest that the sex steroid synthesis pathway may play a role. Glycodelin synthesis is upregulated by progesterone (Seppälä *et al*., 2013), which is upstream of testosterone and oestradiol in the steroid synthesis pathway, with oestradiol regulating SHBG levels. We found little evidence that previously published genetic signals for progesterone levels from the GWAS Catalog were associated with glycodelin (smallest P=0.004, rs112295236 for male glycodelin). The different relationships between sex hormones and glycodelin before and after menopause might reflect the shift from ovarian to adrenal steroidogenesis after menopause. Sex hormones have complex and inter-related effects on signalling pathways within the female reproductive system (Lissaman *et al*., 2023), but the MR methods used should mean that our results reflect direct pathways between the exposure traits tested and glycodelin, i.e. not through correlated traits or reverse causation. Currently there are few robust genetic signals for progesterone and oestradiol, which will be needed to allow sex-hormone pathways to be fully explored using genetic epidemiology. Additionally, in MR genetic instruments are assumed to represent a lifelong exposure, which will not be the case for genetic instruments with different effects before and after menopause and future work should use more sophisticated approaches to more fully investigate pre- and post-menopausal effects.

Apart from sex hormones, we did not find evidence to support glycodelin levels causing, or being caused by, other reproductive conditions other than reproductive lifespan. Later age at menopause raised glycodelin, consistent with observational analyses, where glycodelin levels decline around the average age of menopause (Sun *et al*., 2023). Such changes are consistent with reported changes in endometrial function with ageing (Garcia-Alonso *et al*., 2021). There was some evidence of a relationship between BMI and glycodelin, with higher BMI lowering glycodelin before menopause and raising it after menopause, though no effect of glycodelin on BMI. Previous analysis in UK Biobank (Hillary *et al*., 2024) similarly identified a negative correlation between glycodelin levels and bodyweight before menopause and a positive correlation after menopause, and suggested that genotype at the *PAEP* signal influences this relationship. Despite previously reported relationships of glycodelin with fertility (Suzuki *et al*., 2000) and preeclampsia (Dundar *et al*., 2018), our MR analyses did not find robust evidence to support associations of glycodelin with phenotypes capturing reproductive success, reproductive conditions associated with reduced fertility or preeclampsia. We found some evidence that higher glycodelin levels raised the risk of ovarian cancer, with consistent effects before and after menopause. Higher serum glycodelin levels have been previously associated with less aggressive and benign ovarian tumours (Tsviliana *et al*., 2010; Scholz *et al*., 2012).

Our genome-wide association study identified nine signals associated with glycodelin levels in total, suggesting genes and pathways involved in regulating glycodelin levels in addition to *PAEP*. Two of the signals are pleiotropic for protein levels suggesting overlapping biological pathways: rs1265847 was associated with tumour necrosis factor receptor superfamily member 17 protein, and rs35106244 with both complement receptor type I and glutamate receptor ionotropic kainate protein in UKB*-*PPP. We replicated associations previously reported in the UKB-PPP study (Sun *et al*., 2023), and identified six additional signals passing our genome-wide significance threshold by accounting for sex and menopause status. As would be expected, across all phenotypes the strongest signal was near *PAEP*, the gene encoding glycodelin, which had stronger effects in men than women. The top variant at this signal (rs9409964) is 13kb from *PAEP* but is in an exon of the non-coding transcript *LINC01503*; however, this variant is also an eQTL for *PAEP* in the minor salivary gland (P=3.4×10^-33^). The genetic signal at the *PAEP* locus had a similar absolute effect (in raw units) in pre- and post-menopausal women, which is surprising since *PAEP* is downregulated in the endometrium of ageing women (Devesa-Peiro *et al*., 2022), though such women were 35−45 years and so younger than most women in UK Biobank. Limitations of our analyses were that glycodelin measures were derived from blood and might not reflect levels in reproductive tissues and we were unable to distinguish between the four glycoforms. Protein levels will reflect not only the amount of protein synthesised but also factors such as protein location, relative abundance of important cell types and degradation processes.

Through sex and menopause status stratified analyses, we identified genetic differences in glycodelin regulation, and future releases of proteomic data in UK Biobank will allow these to be replicated. Using our stratified GWAS results in MR analyses allowed us to identify robust causal evidence for a role of testosterone and SHBG in determining glycodelin levels, with differences between men and women and by menopause status. We found little evidence of causal relationships between glycodelin and other reproductive-related traits, in contrast to published associations. In conclusion, we suggest that associations of glycodelin with reproductive conditions are likely to indirectly result from the effects of sex hormones rather than being directly causal.

## Data availability

The genome-wide summary statistics generated by the study will made available in the NHGRI-EBI GWAS Catalog on publication.

## Acknowledgements

The research utilised data from the UK Biobank resource carried out under UK Biobank application number 103356. UK Biobank protocols were approved by the National Research Ethics Service Committee.

## Authors’ roles

S.M. carried out the main analyses. S.M., and K.S.R. drafted the manuscript. S.M., K.S.R, A.M., and J.T. designed the study. All authors were involved in designing and/or performing analysis, revising and approving the manuscript.

## Supplementary Figures

**Supplementary Figure 1:**
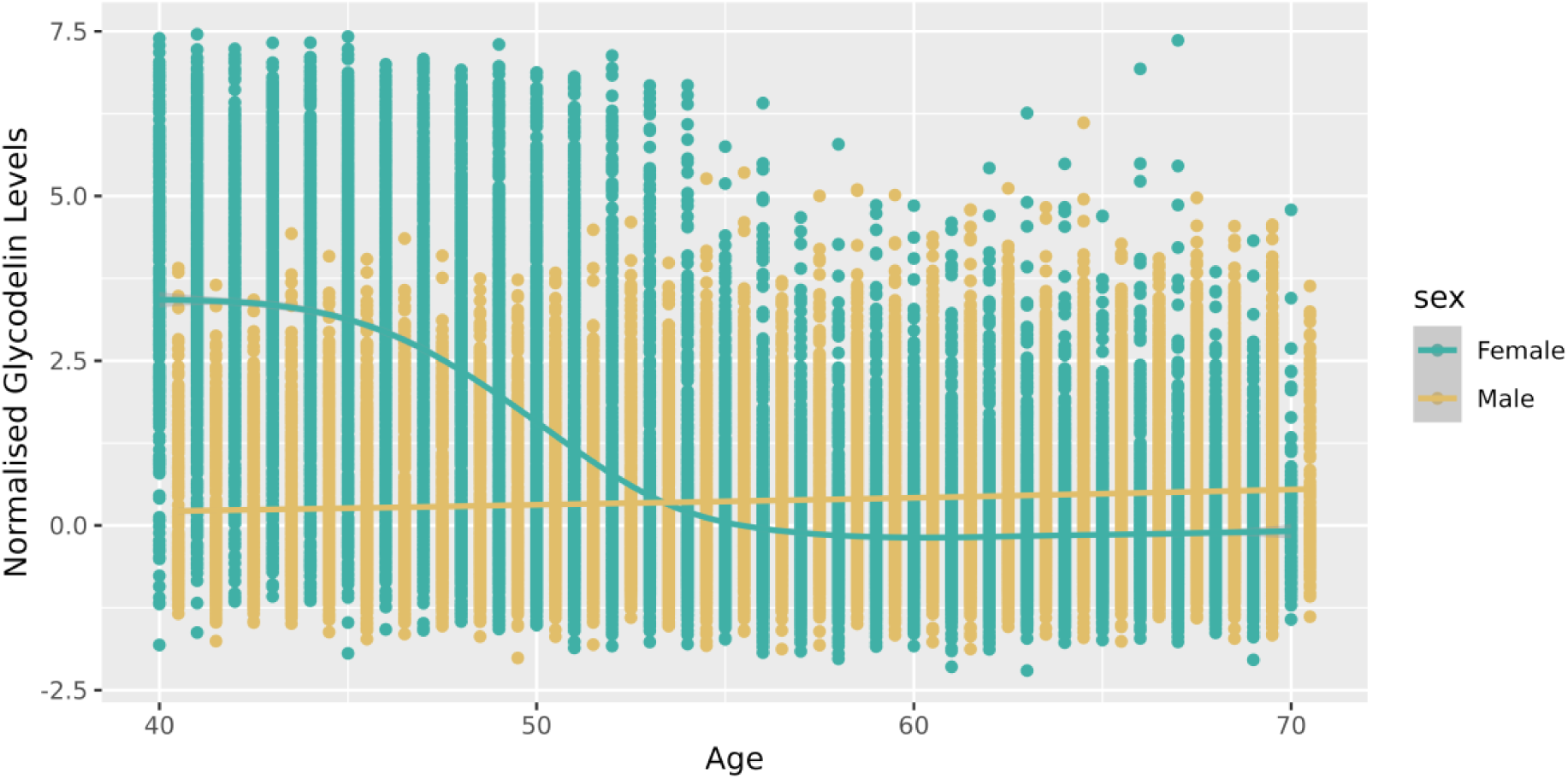
Relationship between glycodelin levels and age in males (n=21,368) and females (n=25,100) in UK Biobank.

**Supplementary Figure 2:**
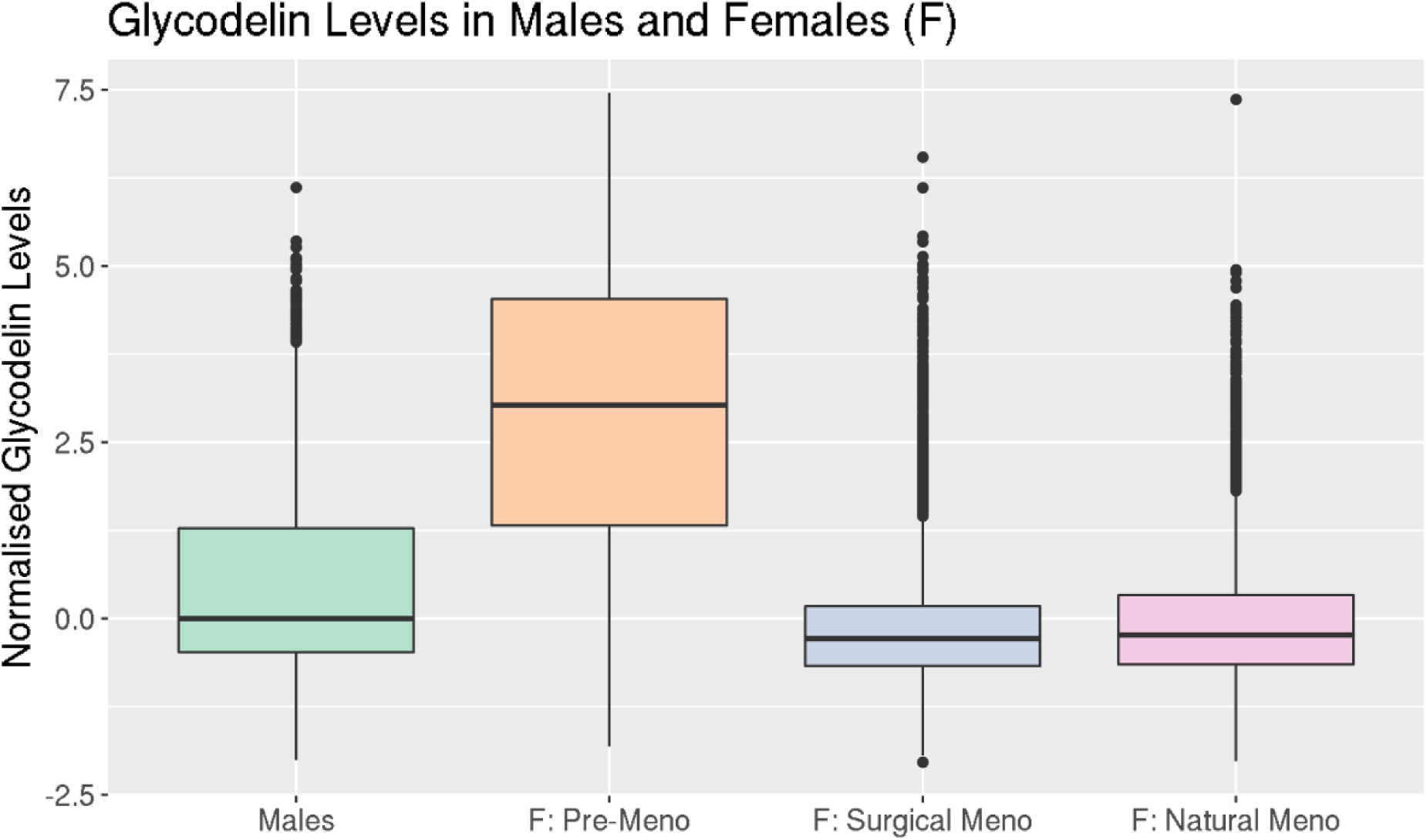
Glycodelin levels (NPX) at the baseline assessment in males, females, as well as pre- and post-menopause. Menopause status was split into natural and surgical (including hysterectomy and bilateral oophorectomy). NPX = normalized protein expression values. Meno = menopause status.

**Supplementary Figure 3:**
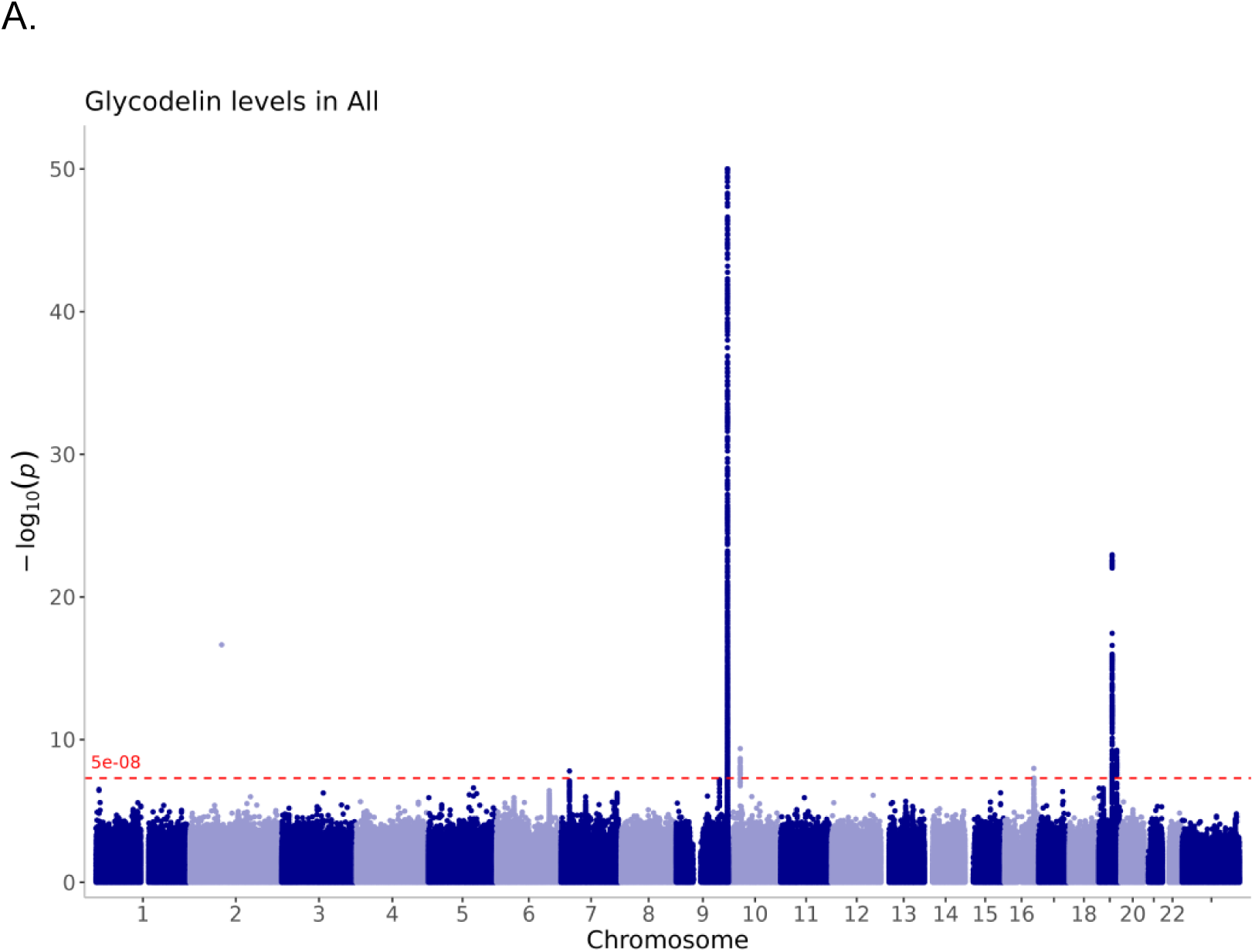
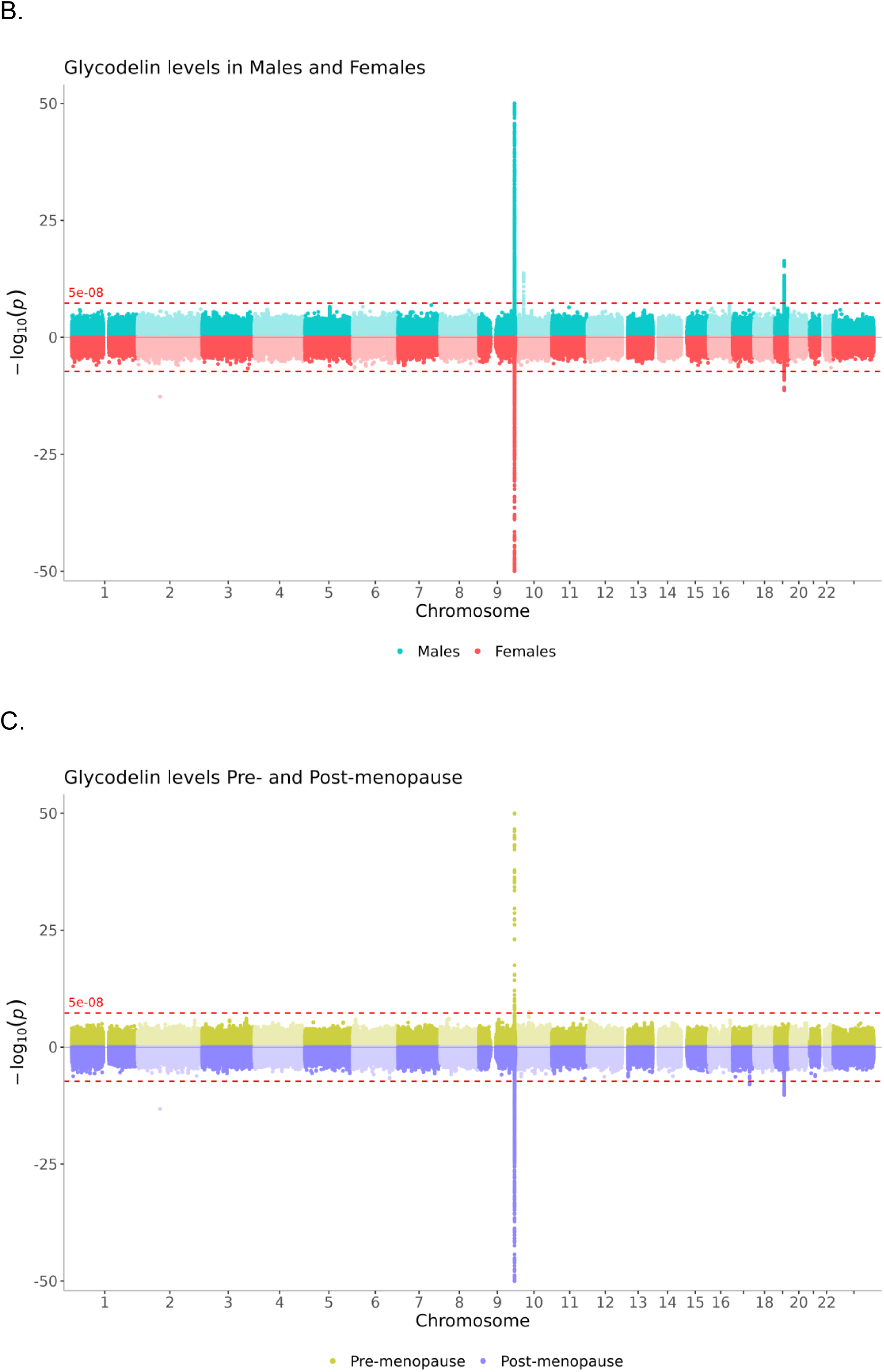
Manhattan plots for the five GWAS conducted of glycodelin. Values with -log10P > 50 were truncated to 50 to reduce the scale. The following plots show: (A) all UKB participants with glycodelin measures (dark blue, n=46,468); (B) males (light blue, n=21,368) and females (red, n=25,100); (C) pre-menopause (green, n=6,409), and post-menopause (purple, n=18,691).

## References

Allen NE, Lacey B, Lawlor DA, Pell JP, Gallacher J, Smeeth L, Elliott P, Matthews PM, Lyons RA, Whetton AD, et al. Prospective study design and data analysis in UK Biobank. Sci Transl Med [Internet] 2024;16:. American Association for the Advancement of Science.

Bowden J, Davey Smith G, Haycock PC, Burgess S. Consistent Estimation in Mendelian Randomization with Some Invalid Instruments Using a Weighted Median Estimator. Genet Epidemiol 2016a;40:304–314. John Wiley & Sons, Ltd.

Bowden J, Del Greco M F, Minelli C, Davey Smith G, Sheehan NA, Thompson JR. Assessing the suitability of summary data for two-sample Mendelian randomization analyses using MR-Egger regression: the role of the I2 statistic. Int J Epidemiol 2016b;45:1961–1974.

Burgess S, Davey Smith G, Davies N, Dudbridge F, Gill D, Glymour M, Hartwig F, Kutalik Z, Holmes M, Minelli C, et al. Guidelines for performing Mendelian randomization investigations: update for summer 2023 [version 3; peer review: 2 approved]. Wellcome Open Res [Internet] 2023;4:.

Burgess S, Labrecque JA. Mendelian randomization with a binary exposure variable: interpretation and presentation of causal estimates. Eur J Epidemiol 2018;33:947–952.

Burney RO, Talbi S, Hamilton AE, Vo KC, Nyegaard M, Nezhat CR, Lessey BA, Giudice LC. Gene Expression Analysis of Endometrium Reveals Progesterone Resistance and Candidate Susceptibility Genes in Women with Endometriosis. Endocrinology 2007;148:3814–3826.

Bycroft C, Freeman C, Petkova D, Band G, Elliott LT, Sharp K, Motyer A, Vukcevic D, Delaneau O, O’Connell J, et al. The UK Biobank resource with deep phenotyping and genomic data. Nature 2018;562:203–209.

Chiu PCN, Chung M-K, Koistinen R, Koistinen H, Seppala M, Ho P-C, Ng EHY, Lee K-F, Yeung WSB. Cumulus Oophorus-associated Glycodelin-C Displaces Sperm-bound Glycodelin-A and -F and Stimulates Spermatozoa-Zona Pellucida Binding *. J Biol Chem 2007;282:5378–5388. Elsevier.

Cui J, Liu Y, Wang X. The Roles of Glycodelin in Cancer Development and Progression. Front Immunol [Internet] 2017;8:.

Davey Smith G, Ebrahim S. ‘Mendelian randomization’: can genetic epidemiology contribute to understanding environmental determinants of disease?*. Int J Epidemiol 2003;32:1–22.

Davies NM, Holmes MV, Davey Smith G. Reading Mendelian randomisation studies: a guide, glossary, and checklist for clinicians. BMJ 2018;362:k601.

Devesa-Peiro A, Sebastian-Leon P, Parraga-Leo A, Pellicer A, Diaz-Gimeno P. Breaking the ageing paradigm in endometrium: endometrial gene expression related to cilia and ageing hallmarks in women over 35 years. Hum Reprod 2022;37:762–776.

Drosdzol-Cop A, Skrzypulec-Plinta V. Selected cytokines and glycodelin A levels in serum and peritoneal fluid in girls with endometriosis. J Obstet Gynaecol Res 2012;38:1245–1253. Australia.

Dundar B, Dincgez Cakmak B, Aydin Boyama B, Karadag B, Ozgen G. Maternal serum glycodelin levels in preeclampsia and its relationship with the severity of the disease. J Matern Fetal Neonatal Med 2018;31:2884–2892. Taylor & Francis.

Garcia-Alonso L, Handfield L-F, Roberts K, Nikolakopoulou K, Fernando RC, Gardner L, Woodhams B, Arutyunyan A, Polanski K, Hoo R, et al. Mapping the temporal and spatial dynamics of the human endometrium in vivo and in vitro. Nat Genet 2021;53:1698–1711.

Hemani G, Zheng J, Elsworth B, Wade KH, Haberland V, Baird D, Laurin C, Burgess S, Bowden J, Langdon R, et al. The MR-Base platform supports systematic causal inference across the human phenome. In Loos R, editor. eLife 2018;7:e34408. eLife Sciences Publications, Ltd.

Hillary RF, Gadd DA, Kuncheva Z, Mangelis T, Lin T, Ferber K, McLaughlin H, Runz H, Marshall E, Marioni RE, et al. Systematic discovery of gene-environment interactions underlying the human plasma proteome in UK Biobank. Nat Commun 2024;15:7346.

Huang L, Rosen JD, Sun Q, Chen J, Wheeler MM, Zhou Y, Min Y-I, Kooperberg C, Conomos MP, Stilp AM, et al. TOP-LD: A tool to explore linkage disequilibrium with TOPMed whole-genome sequence data. Am J Hum Genet 2022;109:1175–1181.

Juliusdottir T. topr: an R package for viewing and annotating genetic association results. BMC Bioinformatics 2023;24:268.

Kocbek V, Vouk K, Mueller MD, Rižner TL, Bersinger NA. Elevated glycodelin-A concentrations in serum and peritoneal fluid of women with ovarian endometriosis. Gynecol Endocrinol 2013;29:455–459. Taylor & Francis.

Lapid K, Sharon N. Meet the multifunctional and sexy glycoforms of glycodelin. Glycobiology 2006;16:39R–45R.

Lissaman AC, Girling JE, Cree LM, Campbell RE, Ponnampalam AP. Androgen signalling in the ovaries and endometrium. Mol Hum Reprod 2023;29:gaad017.

Mbatchou J, Barnard L, Backman J, Marcketta A, Kosmicki JA, Ziyatdinov A, Benner C, O’Dushlaine C, Barber M, Boutkov B, et al. Computationally efficient whole-genome regression for quantitative and binary traits. Nat Genet 2021;53:1097–1103.

McGrath IM, Montgomery GW, Mortlock S, International Endometriosis Genetics Consortium. Polygenic risk score phenome-wide association study reveals an association between endometriosis and testosterone. BMC Med 2023;21:482.

McLaren W, Gil L, Hunt SE, Riat HS, Ritchie GRS, Thormann A, Flicek P, Cunningham F. The Ensembl Variant Effect Predictor. Genome Biol 2016;17:122.

Mosbah A, Nabiel Y, Khashaba E. Interleukin-6, intracellular adhesion molecule-1, and glycodelin A levels in serum and peritoneal fluid as biomarkers for endometriosis. Int J Gynecol Obstet 2016;134:247–251. John Wiley & Sons, Ltd.

Pant A, Moar K, K. Arora T, Maurya PK. Biomarkers of endometriosis. Clin Chim Acta 2023;549:117563.

Purcell S, Neale B, Todd-Brown K, Thomas L, Ferreira MAR, Bender D, Maller J, Sklar P, Bakker PIW de, Daly MJ, et al. PLINK: A Tool Set for Whole-Genome Association and Population-Based Linkage Analyses. Am J Hum Genet 2007;81:559–575.

Ruth KS, Day FR, Tyrrell J, Thompson DJ, Wood AR, Mahajan A, Beaumont RN, Wittemans L, Martin S, Busch AS, et al. Using human genetics to understand the disease impacts of testosterone in men and women. Nat Med 2020;26:252–258.

Ruth KS, Perry JRB, Henley WE, Melzer D, Weedon MN, Murray A. Events in Early Life are Associated with Female Reproductive Ageing: A UK Biobank Study. Sci Rep 2016;6:24710.

Sanderson E, Spiller W, Bowden J. Testing and correcting for weak and pleiotropic instruments in two-sample multivariable Mendelian randomization. Stat Med 2021;40:5434–5452. John Wiley & Sons, Ltd.

Scholz C, Heublein S, Lenhard M, Friese K, Mayr D, Jeschke U. Glycodelin A is a prognostic marker to predict poor outcome in advanced stage ovarian cancer patients. BMC Res Notes 2012;5:551. England.

Seppälä M, Koistinen H, Koistinen R, Chiu PCN, Yeung WSB. Glycodelin: A Lipocalin with Diverse Glycoform-Dependent Actions. Madame Curie Biosci Database Internet [Internet] 2013; Landes Bioscience Available from: https://www.ncbi.nlm.nih.gov/books/NBK6332/.

Seppälä M, Koistinen H, Koistinen R, Hautala L, Chiu PC, Yeung WS. Glycodelin in reproductive endocrinology and hormone-related cancer. Eur J Endocrinol 2009;160:121–133.

Seppälä M, Taylor RN, Koistinen H, Koistinen R, Milgrom E. Glycodelin: A Major Lipocalin Protein of the Reproductive Axis with Diverse Actions in Cell Recognition and Differentiation. Endocr Rev 2002;23:401–430.

Skrivankova VW, Richmond RC, Woolf BAR, Yarmolinsky J, Davies NM, Swanson SA, VanderWeele TJ, Higgins JPT, Timpson NJ, Dimou N, et al. Strengthening the Reporting of Observational Studies in Epidemiology Using Mendelian Randomization: The STROBE-MR Statement. JAMA 2021;326:1614–1621.

Sun BB, Chiou J, Traylor M, Benner C, Hsu Y-H, Richardson TG, Surendran P, Mahajan A, Robins C, Vasquez-Grinnell SG, et al. Plasma proteomic associations with genetics and health in the UK Biobank. Nature 2023;622:329–338.

Suzuki Y, Sugiyama R, Fukumine N, Usuda S, Itoh H, Isaka K, Takayama M, Teisner B. Clinical Applications of Serum Placental Protein 14 (PP14) Measurement in the IVF-ET Cycle. J Obstet Gynaecol Res 2000;26:295–302.

Tsviliana A, Mayr D, Kuhn C, Kunze S, Mylonas I, Jeschke U, Friese K. Determination of Glycodelin-A Expression Correlated to Grading and Staging in Ovarian Carcinoma Tissue. Anticancer Res 2010;30:1637.

Tyrrell J, Mulugeta A, Wood AR, Zhou A, Beaumont RN, Tuke MA, Jones SE, Ruth KS, Yaghootkar H, Sharp S, et al. Using genetics to understand the causal influence of higher BMI on depression. Int J Epidemiol 2019;48:834–848.

Uchida H, Maruyama T, Nishikawa-Uchida S, Miyazaki K, Masuda H, Yoshimura Y. Glycodelin in reproduction. Reprod Med Biol 2013;12:79–84.

Wang P, Zhu L, Zhang X. The role of placental protein 14 in the pathogenesis of endometriosis. Reprod Sci Thousand Oaks Calif 2013;20:1465–1470. United States.

Watanabe K, Taskesen E, Bochoven A van, Posthuma D. Functional mapping and annotation of genetic associations with FUMA. Nat Commun 2017;8:1826.

Yadav S, Kumari P, Sharma S, Aafria S, Batra B, Sharma M. Analytical techniques developed for the determination of glycodelin biomarker: A Mini-Review. Microchem J 2023;195:109394.

